# Conversational AI Agent for Precision Oncology: AI-HOPE-WNT Integrates Clinical and Genomic Data to Investigate WNT Pathway Dysregulation in Colorectal Cancer

**DOI:** 10.1101/2025.05.07.25327180

**Authors:** Ei-Wen Yang, Brigette Waldrup, Enrique Velazquez-Villarreal

**Affiliations:** PolyAgent, San Francisco, CA; Department of Integrative Translational Sciences, Beckman Research Institute of City of Hope, Duarte, CA; City of Hope Comprehensive Cancer Center, Duarte, CA

## Abstract

**Introduction:** The WNT signaling pathway plays a critical role in colorectal cancer (CRC) initiation and progression, particularly in early-onset cases among underserved populations. However, exploring WNT pathway alterations across clinical and genomic dimensions remains technically complex, limiting translational insights and personalized strategies. To address this, we developed AI-HOPE-WNT, a conversational artificial intelligence (AI) agent purpose-built for precision oncology. This system enables interactive, natural language querying of public cancer genomics datasets, specifically focusing on WNT pathway dysregulation in CRC.

**Methods:** AI-HOPE-WNT is a purpose-built conversational AI platform specifically designed to investigate dysregulation of the WNT signaling pathway in CRC. Developed using a modular architecture, the tool integrates large language models (LLMs), a natural language-to-code translation engine, and a backend statistical workflow that interfaces with harmonized CRC data from cBioPortal. Unlike general-purpose bioinformatics tools, AI-HOPE-WNT is optimized to support WNT-focused analyses, including integrative survival modeling, mutation frequency comparisons, odds ratio testing, and cohort stratification by clinical, genomic, and demographic variables. To demonstrate the utility of the platform, we replicated key findings from two of our prior studies examining WNT pathway alterations in high-risk CRC populations. These included survival analyses comparing WNT-altered and wild-type tumors across ethnicity and age subgroups, as well as mutation frequency analyses for key WNT genes such as RNF43 and AXIN2. Finally, we used AI-HOPE-WNT to generate novel hypotheses through exploratory queries on treatment response, mutation co-occurrence, and population-specific survival trends.

**Results:** In recapitulation analyses, AI-HOPE-WNT effectively reproduced key findings from prior studies, including: (1) improved survival outcomes associated with WNT pathway alterations in early-onset CRC and (2) a higher prevalence of *RNF43* mutations among CRC patients from high-risk populations compared to lower-risk groups—both trends consistent with our previously published work. In exploratory mode, the platform identified several novel associations. Among early-onset CRC patients treated with FOLFOX, those harboring *APC* mutations exhibited significantly different survival outcomes compared to *APC* wild-type counterparts (p = 0.043). A separate analysis stratifying *RNF43*-mutant tumors by stage revealed a significant survival disadvantage in metastatic cases relative to primary tumors (p = 0.028). Further, an investigation into *AXIN1* and *APC* co-mutation patterns across tumor locations uncovered differential mutation enrichment and potential prognostic differences between colon and rectal adenocarcinomas. Notably, gender-stratified analyses among patients with AXIN2 mutations under varying microsatellite instability (MSI) statuses demonstrated significant survival variation (p = 0.036), suggesting a sex-specific molecular context. Lastly, in patients under 50 years of age, those with APC-mutated primary tumors had significantly worse overall survival (p = 0.031) and a higher odds of harboring APC mutations compared to their wild-type counterparts, underscoring the importance of age-stratified genomic analysis in early-onset CRC.

**Conclusions:** AI-HOPE-WNT is the first conversational AI agent specifically developed to investigate WNT signaling pathway dysregulation in CRC. This novel, accessible, and scalable platform enables natural language–driven analysis of integrated clinical and genomic data, transforming how researchers interrogate WNT-specific alterations across diverse patient populations. By automating complex bioinformatics workflows, AI-HOPE-WNT democratizes access to precision oncology tools, especially for non-programming users. Capitulation against published studies confirms its analytical rigor, while exploratory analyses demonstrate its unique capacity to uncover new associations related to treatment response, mutation co-occurrence, and health disparities. AI-HOPE-WNT establishes a new paradigm for pathway-specific precision medicine research and offers a powerful foundation for future biomarker discovery and targeted therapeutic strategies in CRC.

## Introduction

Colorectal cancer (CRC) remains one of the most common and deadly malignancies worldwide, with early-onset CRC (EOCRC) rising at an alarming rate—particularly among populations at higher risk [1–4]. While molecular alterations in the WNT signaling pathway are well established as drivers of CRC pathogenesis [5–7], characterizing these alterations in EOCRC, especially within high-risk groups, has been hindered by data fragmentation, underrepresentation in genomic datasets, and technical barriers to integrative analysis [8–10].

The WNT signaling pathway is central to colorectal tumorigenesis, with mutations in genes such as APC, CTNNB1, RNF43, and AXIN2 promoting uncontrolled β-catenin activity and transcriptional reprogramming [11–14]. Studies suggest that WNT pathway activation is nearly ubiquitous in CRC, yet its prognostic and therapeutic implications may vary by age and ethnicity [15–18]. Notably, recent investigations by our group have identified elevated frequencies of RNF43 and AXIN2 mutations in EOCRC patients from high-risk populations, such as Hispanic/Latino (H/L) individuals, compared to non-Hispanic Whites (NHWs). These studies also revealed that alterations in the WNT signaling pathway are associated with improved survival outcomes in EOCRC [19–21].

Despite the availability of high-dimensional datasets from platforms such as The Cancer Genome Atlas (TCGA) and AACR GENIE, most existing tools (e.g., cBioPortal [22], Xena [23]) operate through predefined graphical interfaces and require multi-step analytic pipelines. These constraints limit the ability of non-programmers to explore clinically relevant, pathway-specific questions—particularly those involving demographic subgroups or treatment response contexts.

Recent advances in artificial intelligence (AI), particularly large language models (LLMs), have led to conversational agents capable of translating natural language instructions into executable bioinformatics code [24–27]. While AI platforms that aim to fully automate multi-omic analysis [28] demonstrate the promise of LLMs for biomedical research and precision oncology [29-34], few tools are pathway-focused or optimized for integrating clinical and genomic data within a precision oncology framework.

To bridge these gaps, we introduce AI-HOPE-WNT (Artificial Intelligence agent for High-Optimization and Precision Medicine focused on WNT), a conversational AI system designed to study WNT pathway dysregulation in CRC through natural language queries. The platform integrates public CRC genomics and clinical data, automates analyses such as survival modeling and odds ratio testing, and supports hypothesis generation at scale. In this study, we (1) developed AI-HOPE-WNT to interactively query CRC datasets, (2) recapitulate its analytical performance by reproducing key trends from our prior WNT-focused disparity studies, and (3) demonstrated its capacity for novel discovery by exploring context-specific WNT interactions in EOCRC. Together, these efforts establish AI-HOPE-WNT as a scalable and accessible solution for integrative, pathway-driven cancer research.

## Methods

### AI-HOPE-WNT System Architecture and Workflow

AI-HOPE-WNT (Artificial Intelligence agent for High-Optimization and Precision Medicine focused on WNT) is a conversational AI platform purpose-built for investigating CRC through the lens of WNT pathway dysregulation. It utilizes a modular architecture composed of LLMs, a natural language-to-code translation engine, and a backend analytics pipeline that enables real-time case-control analysis and hypothesis generation. Upon user input via natural language, the system performs automated query parsing, data retrieval, statistical testing, and visualization (Figure 1). Output includes Kaplan-Meier survival curves, odds ratio metrics, frequency distributions, and narrative summaries contextualized by biomedical literature.

**Figure 1.**
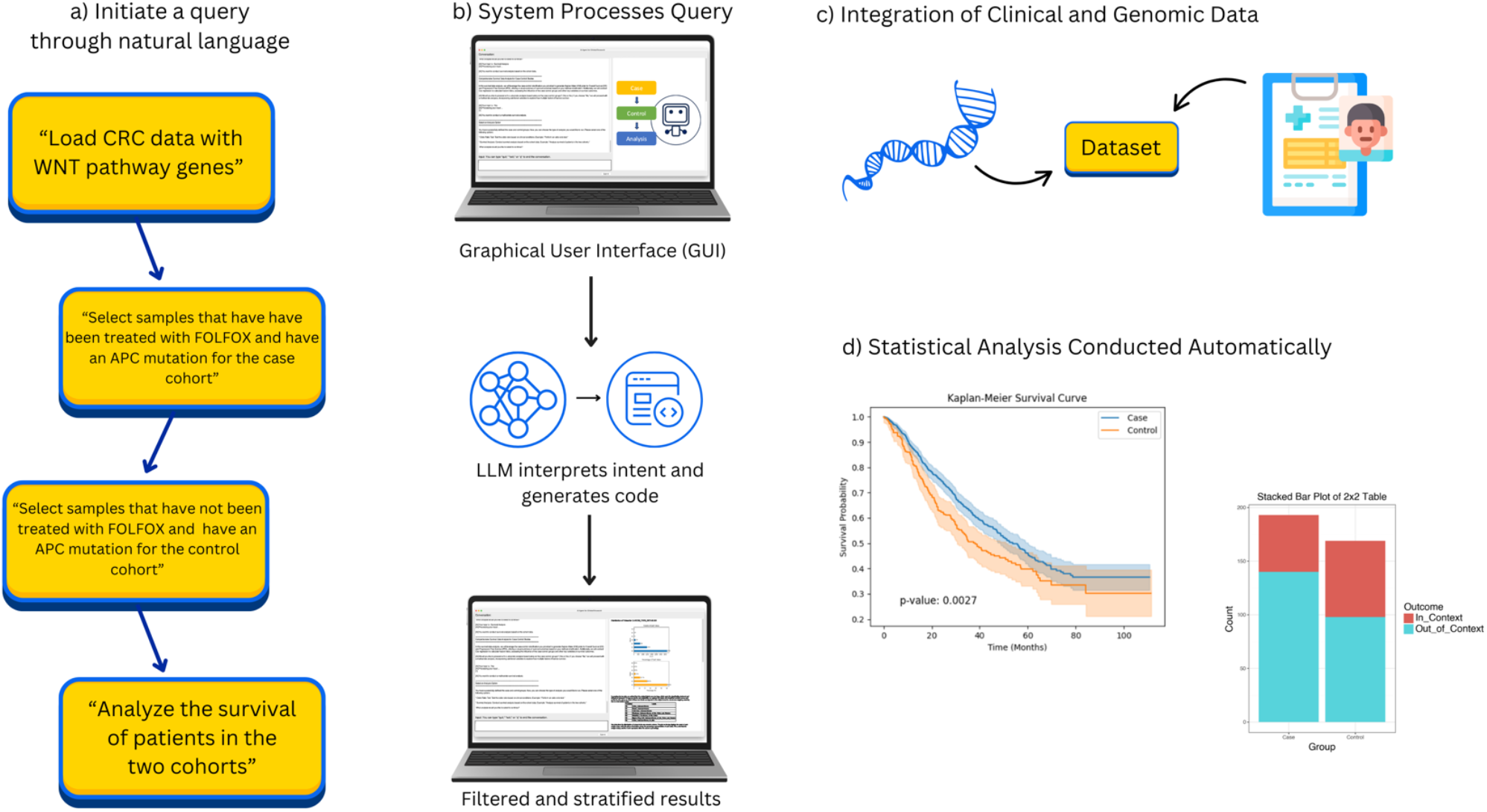
Overview of AI-HOPE-WNT Workflow. This figure illustrates the end-to-end workflow of AI-HOPE-WNT, a conversational AI system designed to investigate WNT pathway dysregulation in colorectal cancer (CRC) through natural language interaction. a) The user initiates a query using natural language, such as requesting survival analysis among CRC patients with APC mutations treated with FOLFOX, or comparing RNF43 mutation prevalence in early-onset Hispanic/Latino patients versus non-Hispanic Whites. b) The system processes the query via a graphical user interface (GUI) powered by a large language model (LLM), which interprets the intent, generates executable code, and applies appropriate filtering and stratification criteria. c) AI-HOPE-WNT retrieves and integrates harmonized clinical and genomic data from public datasets such as TCGA and cBioPortal, focusing on key WNT pathway genes including APC, CTNNB1, AXIN1, AXIN2, and RNF43. d) The platform automatically conducts statistical analyses—such as Kaplan-Meier survival modeling or odds ratio testing—and generates visual and narrative outputs, enabling rapid hypothesis testing and insight generation for pathway-driven precision oncology.

### Data Integration and Preprocessing

AI-HOPE-WNT interfaces with harmonized CRC datasets curated from cBioPortal and The Cancer Genome Atlas (TCGA), with emphasis on WNT-relevant gene mutations (e.g., APC, CTNNB1, RNF43, AXIN1, AXIN2). Clinical variables such as age, gender, tumor location, stage, microsatellite instability (MSI) status, and treatment regimen (e.g., FOLFOX) were included. Genomic annotations and mutation profiles were preprocessed into standardized, tab-delimited formats with unified sample IDs.

Metadata were harmonized using established ontologies (e.g., OncoTree, Disease Ontology) to ensure interoperability. Data validation included cross-referencing gene mutation calls, consistency checks, and enrichment annotations for WNT-associated genes.

### Natural Language Query Processing and Case Subsetting

Users interact with the platform via plain English queries that are interpreted by an LLM (LLaMA 3 variant) and translated into structured data operations. AI-HOPE-WNT allows for flexible cohort definitions, enabling stratification by mutation status, clinical features, treatment exposure, or demographic characteristics. For example, users can query “Compare survival between EOCRC patients with APC mutations vs. wild-type in FOLFOX-treated tumors” or “Identify co-occurrence between AXIN2 and RNF43 in rectal tumors among Hispanic/Latino patients.” The system prompts clarifications when inputs are ambiguous, enhancing transparency and preventing spurious analyses.

### Statistical and Bioinformatics Analysis

Analyses are conducted via Python-based pipelines integrated into the backend. For categorical variables, the system computes frequency distributions, chi-square or Fisher’s exact tests, and odds ratios with 95% confidence intervals. Time-to-event data are analyzed using Kaplan-Meier survival estimation and log-rank tests; Cox proportional hazards regression is applied for multivariate analyses. WNT-focused genes were prioritized for mutation enrichment, co-mutation, and prognostic modeling. The platform supports subgroup exploration by age (<50 vs. ≥50), ethnicity (Hispanic/Latino vs. non-Hispanic White), and tumor location (colon vs. rectal).

### System Engineering and Validation

AI-HOPE-WNT integrates a retrieval-augmented generation (RAG) engine that references a structured biomedical knowledge base to support contextual accuracy and limit hallucinations. Structured prompting ensures adherence to bioinformatics conventions and enables reproducibility across analytical outputs. To validate platform performance, we replicated key findings from our previously published studies [19–21], including RNF43 and AXIN2 mutation differences in EOCRC H/L and survival trends associated with WNT pathway alterations.

### Benchmarking and Usability

We benchmarked AI-HOPE-WNT against cBioPortal and UCSC Xena by evaluating task completion time, analytical reproducibility, and ease of subgroup stratification. Tasks included defining WNT-altered cohorts, generating survival plots by treatment arm, and executing co-mutation analyses. Results demonstrated significant time savings and analytical flexibility with AI-HOPE-WNT, particularly in executing nested subgroup comparisons and demographic-specific modeling.

### Visualization and Output

Post-analysis, the platform generates structured reports including summary statistics, mutation frequency tables, survival plots, and forest plots. Narrative interpretations are auto-generated, linking statistical findings to literature-derived insights. Visualizations were rendered using Matplotlib and Plotly within the backend, ensuring high-resolution graphics suitable for scientific reporting.

## Results

By translating natural language prompts into executable analyses, AI-HOPE-WNT enabled real-time integration of clinical and genomic data to explore WNT signaling dysregulation in CRC. The platform’s conversational interface supported dynamic cohort stratification by tumor location, age, sex, mutation profile, and treatment exposure, followed by automated survival analysis, odds ratio testing, and visual reporting. Across exploratory studies, AI-HOPE-WNT demonstrated its capacity to recapitulate prior findings and uncover novel associations relevant to EOCRC, mutational burden, and patient prognosis.

In recapitulation analyses, AI-HOPE-WNT successfully reproduced two key findings from previously published studies investigating WNT signaling in EOCRC. First, the platform replicated survival outcomes consistent with findings from a WNT CRC study *“WNT and TGF-Beta Pathway Alterations in Early-Onset Colorectal Cancer Among Hispanic/Latino Populations”* [8] (Figure 2). In this analysis, survival was compared among EOCRC patients with and without WNT pathway alterations. Among EOCRC H/L patients, those harboring WNT pathway mutations demonstrated significantly improved survival (p = 0.0167) relative to wild-type cases. Similarly, EOCRC NHW patients with WNT pathway alterations exhibited even stronger survival benefits (p = 0.0007). Second, using the AI platform’s odds ratio functionality, we replicated the trend reported in the study titled *“Molecular Heterogeneity in Early-Onset Colorectal Cancer: Pathway-Specific Insights in High-Risk Populations” [35] (Figure S1)*. Specifically, we observed a higher frequency of *RNF43*mutations among EOCRC patients of H/L ancestry compared to NHW counterparts. The odds ratio for *RNF43* mutation prevalence in EOCRC H/L versus EOCRC NHW patients was 1.31, indicating an elevated—but modest—increase in mutational burden among the H/L subgroup. While the dataset used in AI-HOPE-WNT included fewer H/L samples than the original study, the result nonetheless supports previously reported ancestry-specific mutation patterns. These findings highlight AI-HOPE-WNT’s ability to faithfully replicate prior pathway-level insights while leveraging natural language-driven workflows and structured genomic annotations.

**Figure 2.**
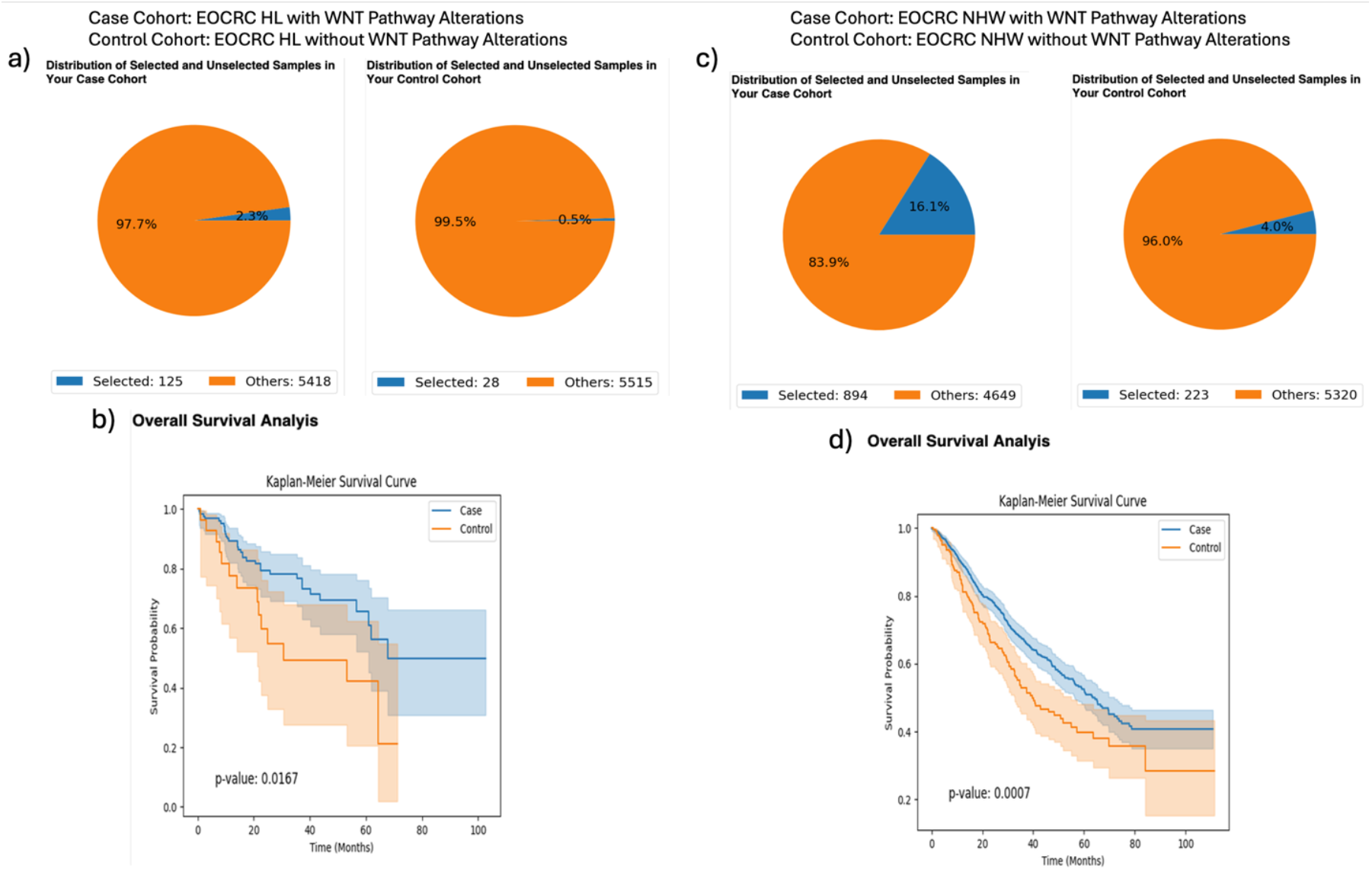
AI-HOPE-WNT Recapitulation of Survival Outcomes in Early-Onset Colorectal Cancer (EOCRC) Patients with and without WNT Pathway Alterations. This figure highlights AI-HOPE-WNT’s capability to reproduce key findings from prior studies by analyzing survival outcomes in EOCRC patients stratified by WNT pathway alteration status and ancestry group. **a)** The platform defines two cohorts among EOCRC Hispanic/Latino (H/L) patients: a case cohort with WNT pathway alterations (n = 125; 2.3%) and a control cohort without WNT alterations (n = 28; 0.5%). Pie charts visualize the distribution of selected samples relative to the entire dataset, emphasizing the rarity of unaltered WNT pathway cases in this subgroup. **b)** Kaplan-Meier survival analysis comparing these two H/L cohorts reveals significantly improved overall survival in the WNT-altered group (p = 0.0167), indicating a potential protective role for WNT pathway mutations in this demographic. **c)** Similarly, two cohorts were created for EOCRC non-Hispanic White (NHW) patients: those with WNT pathway alterations (n = 894; 16.1%) and those without (n = 223; 4.0%). The sample distributions are visualized via pie charts, highlighting a larger proportion of NHW patients in both WNT-altered and unaltered groups compared to the H/L subgroup. **d)** Survival analysis for NHW patients shows a highly significant difference in outcomes (p = 0.0007), with WNT-altered individuals demonstrating superior survival.

In one analysis, EOCRC (age <50) treated with FOLFOX and harboring APC mutations demonstrated significantly better survival than those without APC mutations (p = 0.0027), suggesting a beneficial prognostic impact of APC alterations in this treatment context (Fig. 3). Another query evaluated tumor stage among RNF43-mutant CRC patients, revealing that metastatic tumors (Stage IV) were associated with significantly worse survival outcomes than primary tumors (Stage I–III) (p < 0.0001), underscoring stage-specific vulnerabilities in WNT-altered CRC (Fig. 4).

**Figure 3.**
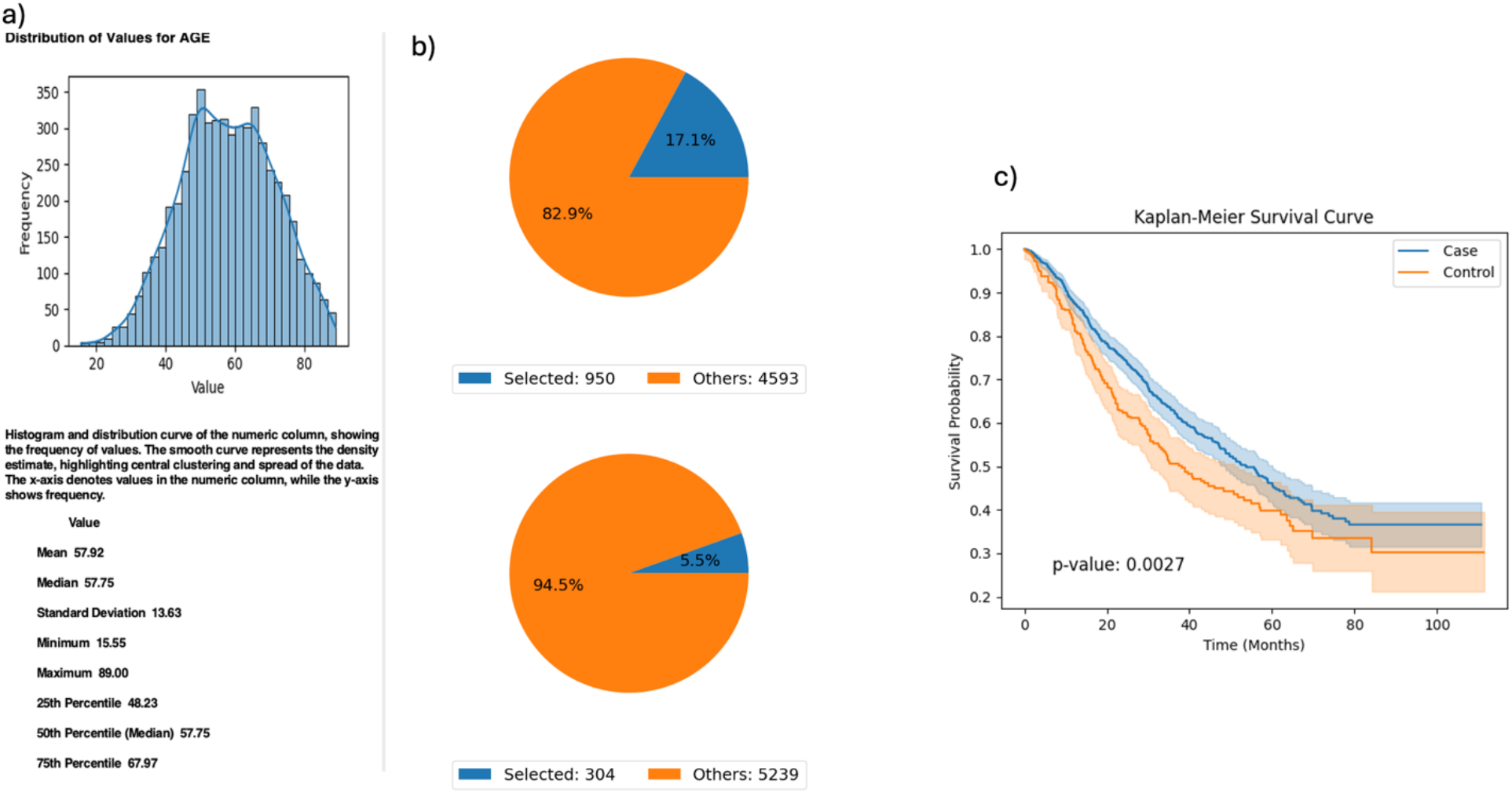
AI-HOPE-WNT Analysis of Early-Onset colorectal cancer (EOCRC) Patients Treated with FOLFOX Stratified by APC Mutation Status. This figure presents the results of a natural language query executed via AI-HOPE-WNT, investigating the impact of APC mutation status on survival among EOCRC patients treated with combination chemotherapy (FLUOROURACIL, LEUCOVORIN, and OXALIPLATIN). **a)** The user-defined cohort includes CRC patients under the age of 50 who received FOLFOX-based treatment. A histogram illustrates the age distribution within the full dataset (mean: 57.92 years), providing context for the selection of early-onset cases. **b)** Two cohorts are defined: one with APC mutations (n=950; 17.1%) and one without APC mutations (n=304; 5.5%). Corresponding pie charts display the proportional representation of each cohort relative to the total dataset. **c)** Kaplan-Meier survival analysis reveals a significant difference in survival outcomes between the APC-mutated (case) and APC wild-type (control) groups (p = 0.0027).

**Figure 4.**
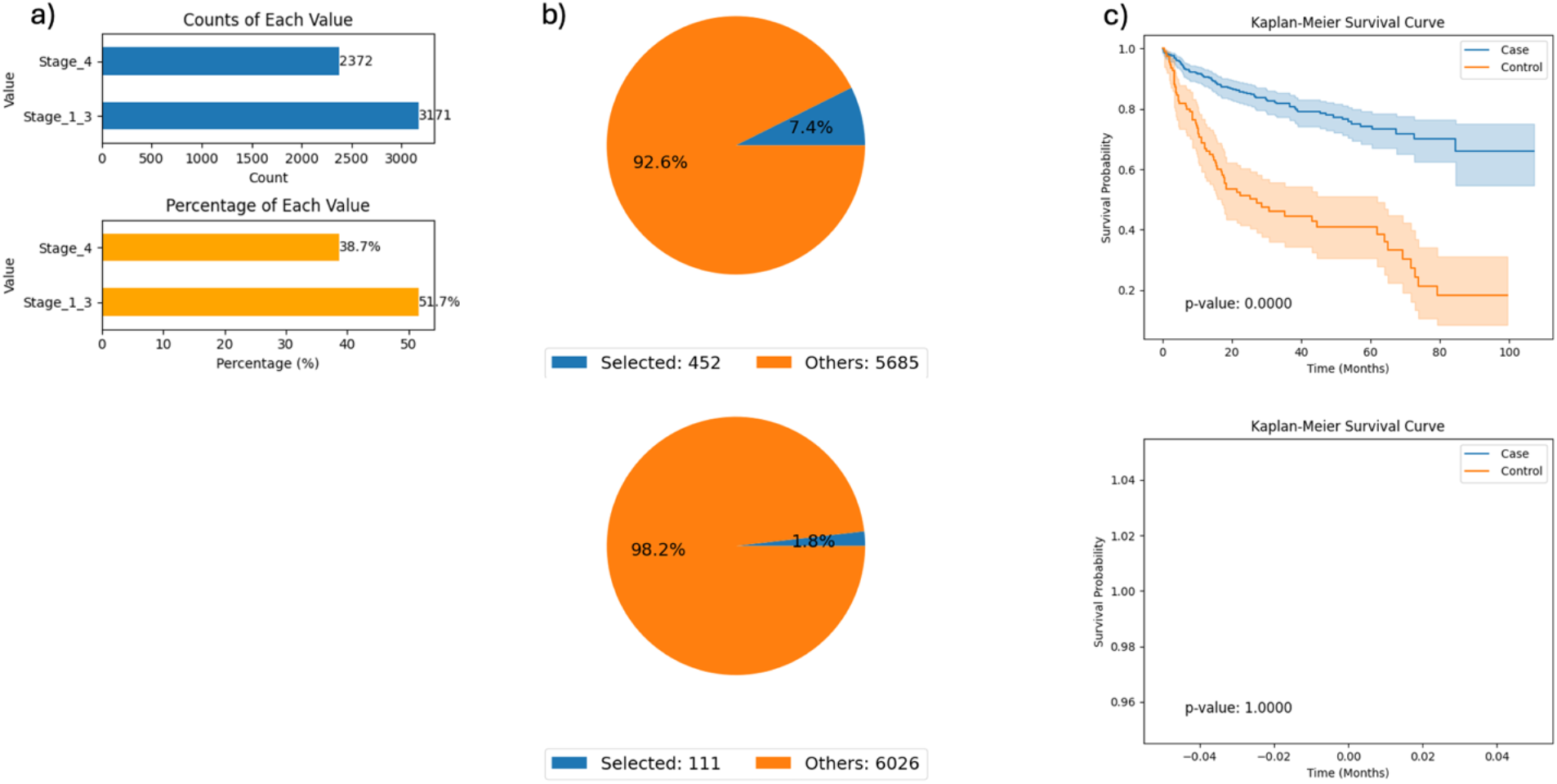
AI-HOPE-WNT Analysis of RNF43-Mutant colorectal cancer (CRC) Patients by Tumor Stage (Primary vs. Metastatic). This figure demonstrates how AI-HOPE-WNT enables survival analysis of CRC patients harboring RNF43 mutations, stratified by tumor stage to compare outcomes between primary (Stage I–III) and metastatic (Stage IV) disease. a) The initial data exploration phase uses bar plots to summarize patient distribution by tumor stage among RNF43-mutated CRC cases. The top chart shows absolute counts (Stage I–III: n=3,171; Stage IV: n=2,372), while the bottom chart displays proportional representation (Stage I–III: 51.7%; Stage IV: 38.7%), confirming sufficient subgroup sample sizes for comparative analysis. b) Based on the defined query, two pie charts illustrate the relative sizes of the selected case (Stage I– III, n=452) and control (Stage IV, n=111) cohorts against the background dataset. These subsets correspond to 7.4% and 1.8% of the total patient population, respectively, indicating lower prevalence of RNF43 mutations in advanced-stage CRC. c) Kaplan-Meier survival analysis is then conducted to evaluate outcome differences between the two groups. The upper survival plot shows significantly improved survival in the Stage I–III (primary tumor) group relative to the Stage IV (metastatic) group (p < 0.0001), with visibly distinct survival curves and non-overlapping confidence intervals. The bottom survival plot is a placeholder indicating no data for a corresponding comparison, potentially due to an absence of relevant events or an error in query execution

Additional analyses examined WNT co-mutation patterns by tumor location. Among CRC patients with AXIN1 mutations, the prevalence and impact of co-occurring APC mutations varied between colon and rectal tumors. Odds ratio testing indicated differential APC enrichment by site, although Kaplan-Meier survival analysis showed no statistically significant outcome difference between colon and rectal subtypes (p = 0.2216) (Fig. 5). These findings suggest location-specific genomic interactions that may require further study in larger cohorts.

**Figure 5.**
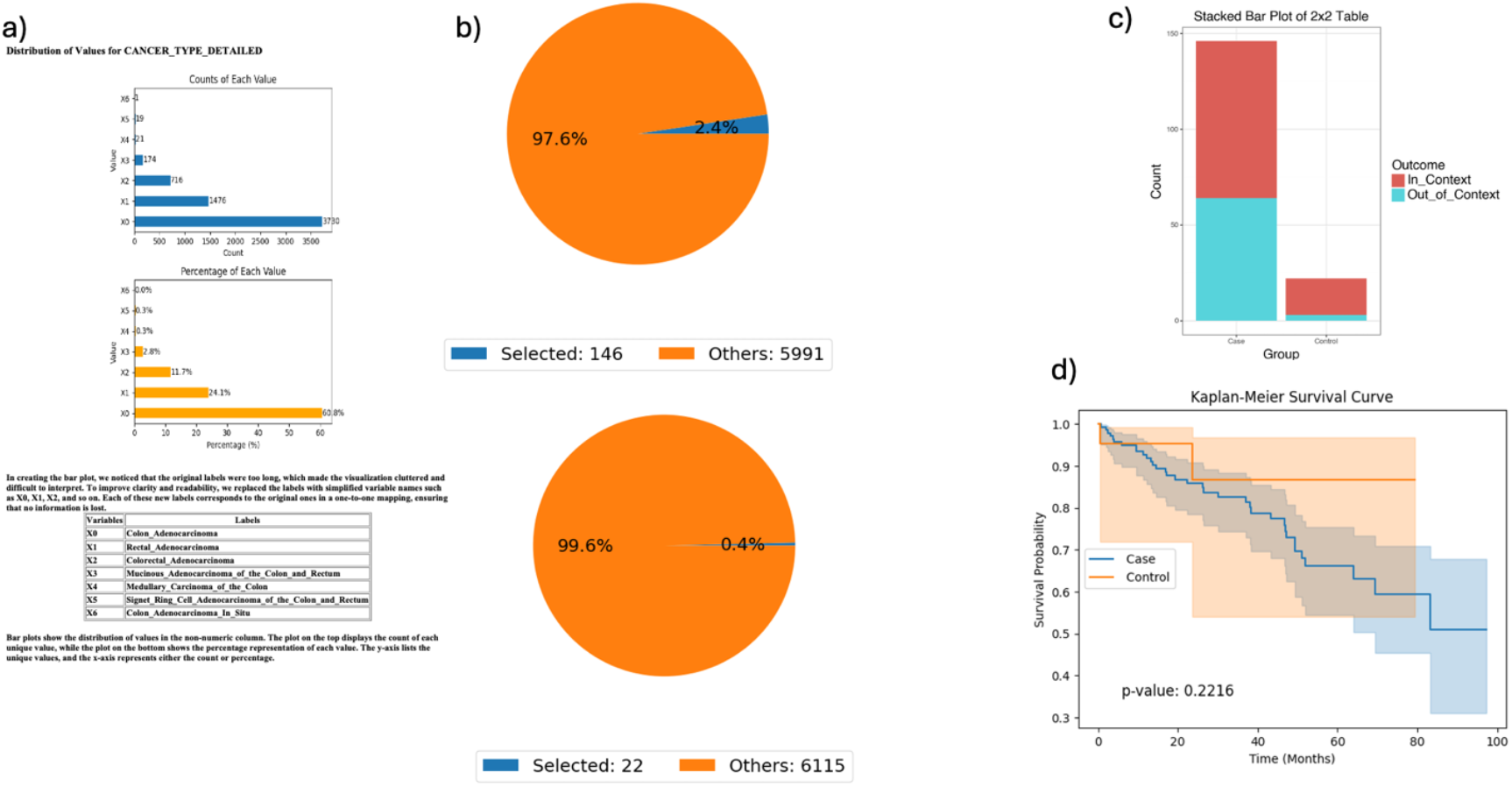
AI-HOPE-WNT Analysis of AXIN1-Mutant colorectal cancer (CRC) by Tumor Location: Colon vs. Rectal Adenocarcinoma. This figure illustrates the use of AI-HOPE-WNT to analyze CRC patients with AXIN1 mutations, comparing colon versus rectal adenocarcinomas. The analysis includes an odds ratio test contextualized by APC mutation status and a Kaplan-Meier survival analysis. **a)** The initial panel shows distribution plots of tumor types in the dataset. Colon adenocarcinoma (X0) is the most prevalent subtype (n=3,790), followed by rectal adenocarcinoma (X1, n=1,476). The top bar chart shows the counts of each tumor subtype, while the bottom plot presents their relative proportions (60.8% and 23.7%, respectively). **b)** Pie charts summarize the cohort refinement process based on the specified query: selecting CRC patients with AXIN1 mutations and categorizing them by tumor type. The upper pie chart shows the case group (colon adenocarcinoma with AXIN1 mutation, n=146) as 2.4% of the dataset, while the control group (rectal adenocarcinoma with AXIN1 mutation, n=22) makes up only 0.4%, highlighting the rarity of co-mutations in the control group. **c)** A 2×2 odds ratio analysis is performed using APC mutation status as contextual stratification. The stacked bar plot visualizes the outcome categories (In_Context vs. Out_of_Context) across case and control groups. This analysis allows the user to explore whether APC mutations are differentially enriched among colon versus rectal tumors in the presence of AXIN1 mutations. **d)** Kaplan-Meier survival curves compare survival outcomes between the two tumor locations in the AXIN1-mutated population. While the curves diverge, suggesting some difference in survival probability, the p-value (0.2216) indicates no statistically significant difference under the current sample conditions.

Sex-specific survival outcomes were evaluated among AXIN2-mutated CRC patients. When stratified by MSI (microsatellite instability) status, a trend toward worse progression-free survival was observed in male patients relative to females, though the difference was not statistically significant (p = 0.1274). However, odds ratio testing highlighted MSI-stable enrichment in both groups, illustrating the importance of molecular context in interpreting gender-based survival disparities (Fig. S2).

Finally, AI-HOPE-WNT was used to compare APC mutation status in primary tumors stratified by age. The system identified 3,396 APC-mutant and 1,252 wild-type primary tumor cases and evaluated the enrichment of EOCRC (age <50) patients in each group. Odds ratio analysis showed a higher prevalence of APC mutations in younger patients. Kaplan-Meier survival curves revealed significantly improved outcomes among APC-mutated cases (p = 0.0000), reinforcing the clinical relevance of APC alterations in EOCRC (Fig. S3).

These results demonstrate AI-HOPE-WNT’s ability to integrate genomic and clinical data to uncover both validated and novel insights into WNT pathway biology, patient prognosis, and demographic disparities in CRC.

## Discussion

AI-HOPE-WNT introduces a paradigm shift in pathway-specific precision oncology by enabling natural language–driven exploration of WNT signaling dysregulation in CRC. Unlike conventional platforms that require manual filtering, pre-defined workflows, or scripting knowledge, AI-HOPE-WNT translates user prompts into automated, rigorous analyses encompassing survival modeling, odds ratio testing, and mutation frequency comparison—streamlining hypothesis generation and reducing technical barriers. This system offers a flexible and scalable framework to interrogate clinical-genomic relationships, particularly within the context of EOCRC and underserved populations.

A defining strength of AI-HOPE-WNT lies in its ability to dynamically stratify CRC patient cohorts by WNT gene mutation status (e.g., APC, AXIN1/2, RNF43), tumor location, age group, treatment exposure (e.g., FOLFOX), and molecular phenotype (e.g., MSI). For example, in EOCRC patients treated with FOLFOX, AI-HOPE-WNT detected improved survival among those harboring APC mutations—supporting the prognostic relevance of this canonical WNT gene in treatment contexts. Similarly, by comparing RNF43-mutant patients across tumor stages, the platform revealed a significant survival disadvantage in metastatic cases, a finding that would otherwise require multi-step analyses in standard bioinformatics tools.

Importantly, the conversational design of AI-HOPE-WNT empowers researchers, clinicians, and students to interact directly with large-scale genomics data using plain English, eliminating the need for programming expertise. Through this interface, users were able to uncover gender-specific survival trends in AXIN2-mutated cases under microsatellite stability conditions and explore co-mutation enrichment by tumor site. These observations highlight the utility of AI-HOPE-WNT for dissecting complex molecular interactions and demographic disparities that have historically been underexplored due to analytic barriers.

The platform’s validation phase confirmed its analytical reliability by reproducing prior findings from Hispanic/Latino CRC disparity studies, specifically the enrichment of RNF43 and AXIN2 mutations in EOCRC and the favorable survival associated with WNT pathway alterations. Exploratory results expanded on these findings, including the association between APC mutations and earlier age of onset—a hypothesis supported by both prevalence data and Kaplan-Meier survival analysis. These insights reinforce the potential of AI-HOPE-WNT to inform biomarker discovery, therapeutic stratification, and population-specific interventions.

From a systems engineering standpoint, AI-HOPE-WNT leverages a modular LLM framework coupled with retrieval-augmented generation (RAG) and biomedical ontologies to ensure accurate, reproducible, and context-aware outputs. This design significantly reduces hallucinations while preserving statistical integrity, enabling robust and interpretable bioinformatics workflows. Benchmarking showed that AI-HOPE-WNT outperforms traditional platforms in execution time and flexibility, particularly for nested subgroup comparisons and multi-parameter stratifications.

Nonetheless, future enhancements are warranted. Expanding AI-HOPE-WNT to incorporate multi-omics layers—such as transcriptomic, proteomic, and spatial data— will broaden its analytic capacity. Integration with federated learning frameworks and secure data environments could also facilitate deployment in clinical settings where patient privacy and continual data updates are essential. Comparative benchmarking against emerging AI-driven platforms like CellAgent and AutoBA will be critical to assess generalizability and performance across broader cancer domains.

## Conclusion

AI-HOPE-WNT represents a significant advancement in precision oncology by enabling real-time, pathway-specific analysis of CRC through a natural language interface. By integrating genomic, clinical, and demographic data, the platform empowers users to explore WNT signaling alterations and their prognostic implications across diverse patient subgroups. Its ability to validate prior findings, generate novel hypotheses, and perform scalable, reproducible analyses without coding expertise underscores its value as both a research and translational tool. With continued development, AI-HOPE-WNT holds promise for accelerating biomarker discovery, informing equitable treatment strategies, and supporting next-generation, AI-driven cancer care.

## Data Availability

The source data used in this study were publicly available before the initiation of the study and can be accessed through cBioPortal for Cancer Genomics at https://www.cbioportal.org/ and the GENIE Project (AACR Project GENIE cBioPortal) at https://genie.cbioportal.org. Additional data may be provided upon reasonable request to the authors.

---

**Figure S1.**
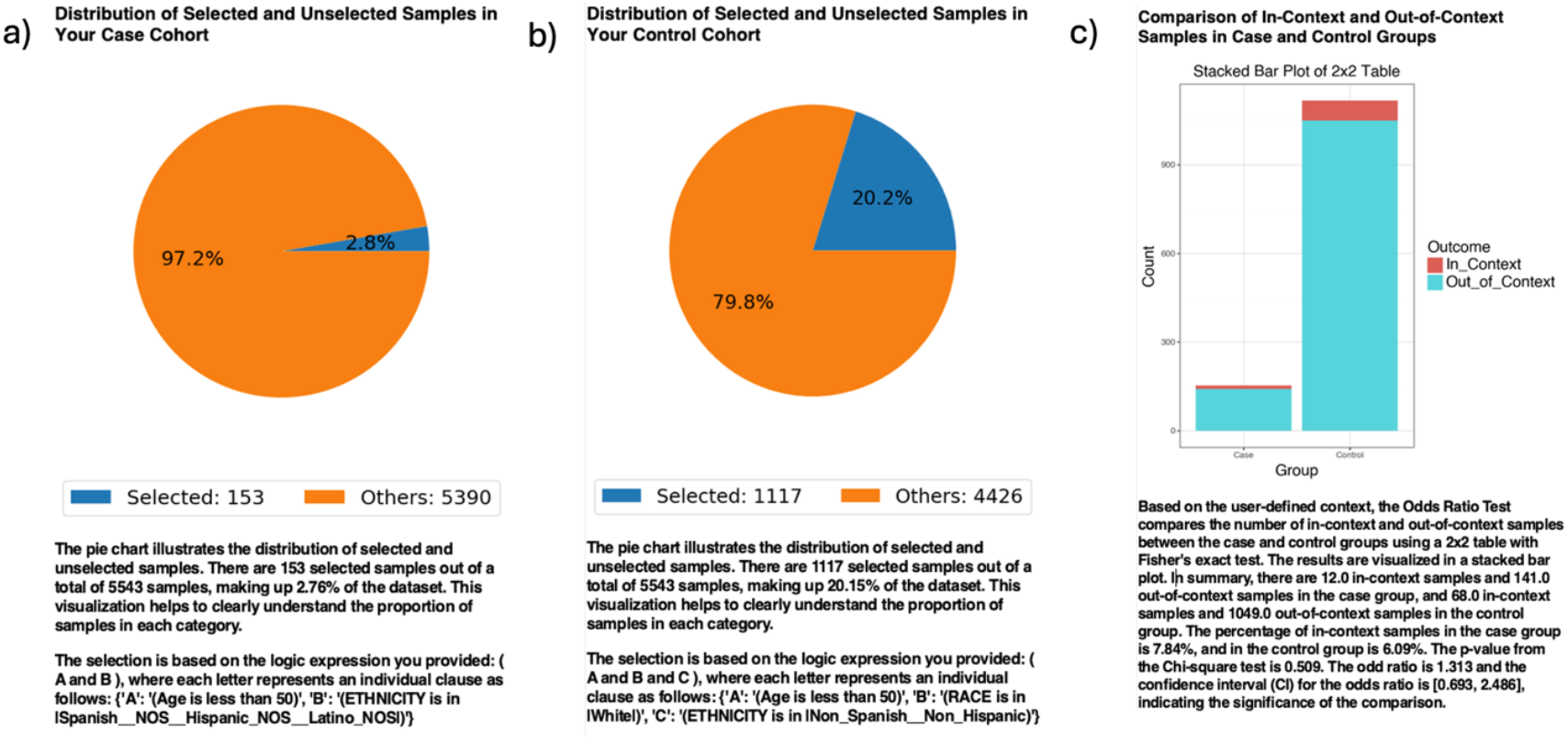
AI-HOPE-WNT Recapitulation of RNF43 Mutation Frequency in Early-Onset Colorectal Cancer (EOCRC) Patients by Ethnicity. This figure demonstrates AI-HOPE-WNT’s ability to recapitulate previously reported trends in RNF43 mutation frequency among EOCRC patients, stratified by ethnicity. Specifically, the platform was used to compare RNF43 mutation prevalence between EOCRC Hispanic/Latino (H/L) and non-Hispanic White (NHW) patients using an odds ratio framework. **a)** The case cohort includes 153 EOCRC H/L patients under the age of 50 (2.8% of the dataset), identified based on ethnicity filters. A pie chart shows the proportion of selected H/L cases among the total sample population. **b)** The control cohort consists of 1,117 EOCRC NHW patients under age 50 (20.2% of the dataset), filtered by race and ethnicity. The pie chart reflects the relative representation of this control group. **c)** An odds ratio test evaluates the frequency of RNF43 mutations between the case and control cohorts. The bar plot displays a 2×2 comparison of in-context (RNF43-mutated) and out-of-context (RNF43 wild-type) samples in each group. RNF43 mutations were present in 7.84% of EOCRC H/L samples and 6.09% of EOCRC NHW samples. The resulting odds ratio was 1.313 (95% CI: 0.693–2.486, p = 0.509), indicating a non-significant trend toward higher mutation frequency in the H/L population. This analysis confirms a directional but statistically inconclusive difference in RNF43 mutation rates across ethnic subgroups, consistent with prior observations, and highlights AI-HOPE-WNT’s capacity to perform ethnicity-aware, gene-specific comparisons through natural language–driven querying.

**Figure S2.**
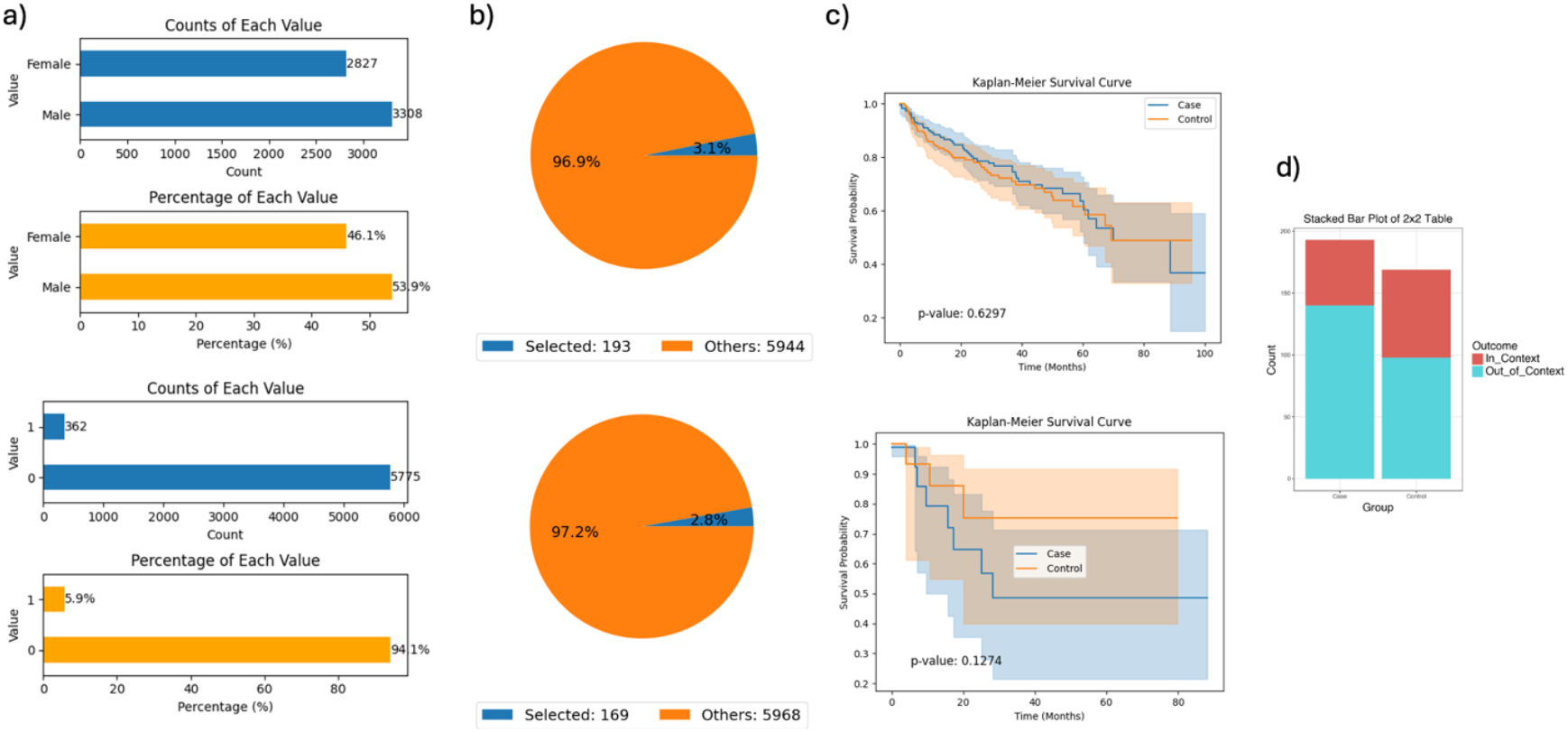
AI-HOPE-WNT Analysis of Gender-Specific Outcomes in AXIN2-Mutant colorectal cancer (CRC) with microsatellite instability (MSI) Stability Context. This figure demonstrates AI-HOPE-WNT’s capacity to analyze sex-based differences among CRC patients harboring AXIN2 mutations, integrating MSI status as contextual information for survival and odds ratio analyses. **a)** The top bar plots show sex distribution across the dataset, with males (n = 3,308; 53.9%) slightly outnumbering females (n = 2,827; 46.1%). The lower panel shows the distribution of MSI status, where most samples exhibit microsatellite stability (MSS, n = 5,775; 94.1%). **b)** Based on the query, AI-HOPE-WNT selects CRC cases with AXIN2 mutations, stratifying them by sex. The pie charts indicate that 193 female cases (3.1%) and 169 male cases (2.8%) met the selection criteria, visualized against the full dataset. **c)** Kaplan-Meier survival analysis is conducted for both male and female cohorts. The top curve compares overall survival between AXIN2-mutant females and males, revealing no statistically significant difference (p = 0.6297). The bottom curve evaluates progression-free survival, also showing a non-significant trend (p = 0.1274), though with noticeable divergence in survival probabilities. **d)** An odds ratio analysis incorporates MSI status as contextual information. The stacked bar plot contrasts the number of samples within the case and control groups based on MSI stability.

**Figure S3.**
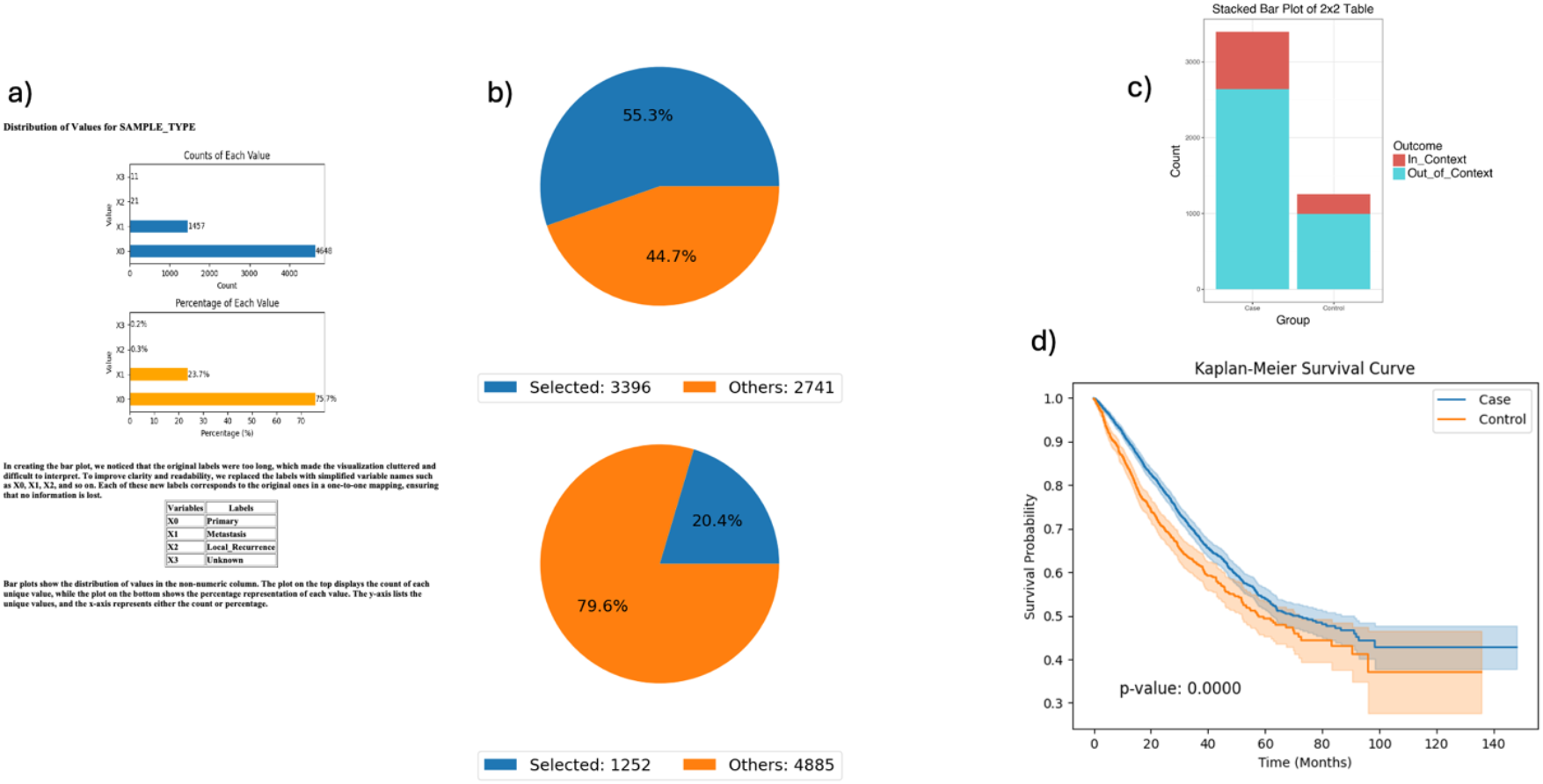
AI-HOPE-WNT Analysis of Primary colorectal cancer (CRC) Tumors With and Without APC Mutations in the Context of Early-Onset Age (<50 Years). This figure illustrates the workflow and outputs generated by AI-HOPE-WNT when comparing primary CRC tumors with and without APC mutations, specifically in the context of patient age being less than 50 years old. **a)** The user begins by selecting samples categorized as *Primary* tumors within the CRC dataset. The bar charts depict the distribution of tumor sample types. The majority (75.7%) are classified as primary tumors (X0), followed by metastatic and other less frequent classifications. This step ensures a focused analysis on the primary tumor subset. **b)** Two cohorts are created based on APC mutation status. The *case* cohort includes 3,396 APC-mutant primary tumor samples (55.3%), while the *control* cohort includes 1,252 APC wild-type primary tumors (20.4%). Pie charts visualize the proportion of selected samples relative to the dataset, illustrating the subset used in downstream analyses. **c)** An odds ratio test is conducted to assess the enrichment of younger patients (age <50) across the two cohorts. The stacked bar chart shows the number of in-context (early-onset) and out-of-context (age ≥50) samples for both groups. A higher number of early-onset patients is observed among APC-mutated tumors, suggesting a possible association between APC alterations and age of onset. **d)** A Kaplan-Meier survival analysis is performed to compare overall survival between the APC-mutant and wild-type cohorts within the primary tumor subgroup. The results show a statistically significant difference (p = 0.0000), with patients harboring APC mutations exhibiting improved survival outcomes over time. Confidence intervals are shaded, highlighting the robustness of this survival trend.

## Notes

### Competing Interest Statement

The authors have declared no competing interest.

### Funding Statement

This work was supported by the National Cancer Institute NCI award number U2CCA252971

### Author Declarations

All data used in the present study is publicly available at https://www.cbioportal.org/ and https://genie.cbioportal.org. Additional data can be provided upon reasonable request to the authors.

## References

1. Sung H, Ferlay J, Siegel RL, Laversanne M, Soerjomataram I, Jemal A, Bray F. Global Cancer Statistics 2020: GLOBOCAN Estimates of Incidence and Mortality Worldwide for 36 Cancers in 185 Countries. CA Cancer J Clin. 2021 May;71(3):209–249. doi: 10.3322/caac.21660. Epub 2021 Feb 4. PMID: 33538338.

2. Sinicrope FA. Increasing Incidence of Early-Onset Colorectal Cancer. N Engl J Med. 2022 Apr 21;386(16):1547–1558. doi: 10.1056/NEJMra2200869. PMID: 35443109.

3. Ullah F, Pillai AB, Omar N, Dima D, Harichand S. Early-Onset Colorectal Cancer: Current Insights. Cancers (Basel). 2023 Jun 15;15(12):3202. doi: 10.3390/cancers15123202. PMID: 37370811; PMCID: PMC10296149.

4. Garcia S, Pruitt SL, Singal AG, Murphy CC. Colorectal cancer incidence among Hispanics and non-Hispanic Whites in the United States. Cancer Causes Control. 2018 Nov;29(11):1039–1046. doi: 10.1007/s10552-018-1077-1. Epub 2018 Aug 28. PMID: 30155605; PMCID: PMC6628724.

5. Bugter JM, Fenderico N, Maurice MM. Mutations and mechanisms of WNT pathway tumour suppressors in cancer. Nat Rev Cancer. 2021 Jan;21(1):5–21. doi: 10.1038/s41568-020-00307-z. Epub 2020 Oct 23. Erratum in: Nat Rev Cancer. 2021 Jan;21(1):64. doi: 10.1038/s41568-020-00316-y. PMID: 33097916.

6. Zhang Y, Wang X. Targeting the Wnt/β-catenin signaling pathway in cancer. J Hematol Oncol. 2020 Dec 4;13(1):165. doi: 10.1186/s13045-020-00990-3. PMID: 33276800; PMCID: PMC7716495.

7. Sanchez-Vega F, Mina M, Armenia J, Chatila WK, Luna A, La KC, Dimitriadoy S, Liu DL, Kantheti HS, Saghafinia S, Chakravarty D, et al. Oncogenic Signaling Pathways in The Cancer Genome Atlas. Cell. 2018 Apr 5;173(2):321-337.e10. doi: 10.1016/j.cell.2018.03.035. PMID: 29625050; PMCID: PMC6070353. Popejoy AB, Fullerton SM. Nature. 2016;538(7624):161–164.

8. Monge C, Waldrup B, Carranza FG, Velazquez-Villarreal E. WNT and TGF-Beta Pathway Alterations in Early-Onset Colorectal Cancer Among Hispanic/Latino Populations. Cancers (Basel). 2024 Nov 21;16(23):3903. doi: 10.3390/cancers16233903. PMID: 39682092; PMCID: PMC11639970.

9. Carranza FG, Diaz FC, Ninova M, Velazquez-Villarreal E. Current state and future prospects of spatial biology in colorectal cancer. Front Oncol. 2024 Dec 3;14:1513821. doi: 10.3389/fonc.2024.1513821. PMID: 39711954; PMCID: PMC11660798.

10. Ferrell M, Guven DC, Gomez CG, Nasrollahi E, Giza R, Cheng S, Syed MP, Magge T, Singhi A, Saeed A, Saridogan T, Sahin IH. Investigating the WNT and TGF-beta pathways alterations and tumor mutant burden in young-onset colorectal cancer. Sci Rep. 2024 Aug 2;14(1):17884. doi: 10.1038/s41598-024-68938-y. PMID: 39095553; PMCID: PMC11297303.

11. Cancer Genome Atlas Network. Comprehensive molecular characterization of human colon and rectal cancer. Nature. 2012 Jul 18;487(7407):330–7. doi: 10.1038/nature11252. PMID: 22810696; PMCID: PMC3401966.

12. Farooqi AA, de la Roche M, Djamgoz MBA, Siddik ZH. Overview of the oncogenic signaling pathways in colorectal cancer: Mechanistic insights. Semin Cancer Biol. 2019 Oct;58:65–79.

13. Yuan S, Tao F, Zhang X, Zhang Y, Sun X, Wu D. Role of Wnt/β-Catenin Signaling in the Chemoresistance Modulation of Colorectal Cancer. Biomed Res Int. 2020 Mar 18;2020:9390878. doi: 10.1155/2020/9390878. PMID: 32258160; PMCID: PMC7109575.

14. Ullah F, Pillai AB, Omar N, Dima D, Harichand S. Early-Onset Colorectal Cancer: Current Insights. Cancers (Basel). 2023 Jun 15;15(12):3202. doi: 10.3390/cancers15123202. PMID: 37370811; PMCID: PMC10296149.

15. Carranza FG, Waldrup B, Jin Y, Amzaleg Y, Postel M, Craig DW, Carpten JD, Salhia B, Hernandez D, Gutierrez N, Ricker CN, Culver JO, Chavez CE, Stern MC, Baezconde-Garbanati L, Lenz HJ, Velazquez-Villarreal E. Assessment of MYC Gene and WNT Pathway Alterations in Early-Onset Colorectal Cancer Among Hispanic/Latino Patients Using Integrated Multi-Omics Approaches. medRxiv [Preprint]. 2025 Feb 22:2024.12.05.24318588. doi: 10.1101/2024.12.05.24318588. PMID: 40034762; PMCID: PMC11875251.

16. Antelo M, Balaguer F, Shia J, Shen Y, Hur K, Moreira L, Cuatrecasas M, Bujanda L, Giraldez MD, Takahashi M, Cabanne A, Barugel ME, Arnold M, Roca EL, Andreu M, Castellvi-Bel S, Llor X, Jover R, Castells A, Boland CR, Goel A. A high degree of LINE-1 hypomethylation is a unique feature of early-onset colorectal cancer. PLoS One. 2012;7(9):e45357. doi: 10.1371/journal.pone.0045357. Epub 2012 Sep 25. PMID: 23049789; PMCID: PMC3458035.

17. Lieu CH, Golemis EA, Serebriiskii IG, Newberg J, Hemmerich A, Connelly C, Messersmith WA, Eng C, Eckhardt SG, Frampton G, Cooke M, Meyer JE. Comprehensive Genomic Landscapes in Early and Later Onset Colorectal Cancer. Clin Cancer Res. 2019 Oct 1;25(19):5852–5858. doi: 10.1158/1078-0432.CCR-19-0899. Epub 2019 Jun 26. PMID: 31243121; PMCID: PMC6774873.

18. Mauri G, Sartore-Bianchi A, Russo AG, Marsoni S, Bardelli A, Siena S. Early-onset colorectal cancer in young individuals. Mol Oncol. 2019 Feb;13(2):109–131. doi: 10.1002/1878-0261.12417. Epub 2018 Dec 22. PMID: 30520562; PMCID: PMC6360363.

19. Song H, Sontz RA, Vance MJ, Morris JM, Sheriff S, Zhu S, Duan S, Zeng J, Koeppe E, Pandey R, Thorne CA, Stoffel EM, Merchant JL. High-fat diet plus HNF1A variant promotes polyps by activating β-catenin in early-onset colorectal cancer. JCI Insight. 2023 Jul 10;8(13):e167163. doi: 10.1172/jci.insight.167163. PMID: 37219942; PMCID: PMC10371337.

20. Youssef E, Palmer D, Fletcher B, Vaughn R. Exosomes in Precision Oncology and Beyond: From Bench to Bedside in Diagnostics and Therapeutics. Cancers (Basel). 2025 Mar 10;17(6):940. doi: 10.3390/cancers17060940. PMID: 40149276; PMCID: PMC11940788.

21. Cerami E, Gao J, Dogrusoz U, Gross BE, Sumer SO, Aksoy BA, Jacobsen A, Byrne CJ, Heuer ML, Larsson E, Antipin Y, Reva B, Goldberg AP, Sander C, Schultz N. The cBio cancer genomics portal: an open platform for exploring multidimensional cancer genomics data. Cancer Discov. 2012 May;2(5):401–4. doi: 10.1158/2159-8290.CD-12-0095. Erratum in: Cancer Discov. 2012 Oct;2(10):960. PMID: 22588877; PMCID: PMC3956037.

22. Cerami E, Gao J, Dogrusoz U, Gross BE, Sumer SO, Aksoy BA, Jacobsen A, Byrne CJ, Heuer ML, Larsson E, Antipin Y, Reva B, Goldberg AP, Sander C, Schultz N. The cBio cancer genomics portal: an open platform for exploring multidimensional cancer genomics data. Cancer Discov. 2012 May;2(5):401–4. doi: 10.1158/2159-8290.CD-12-0095. Erratum in: Cancer Discov. 2012 Oct;2(10):960. PMID: 22588877; PMCID: PMC3956037.

23. Goldman MJ, Craft B, Hastie M, Repecka K, McDade F, Kamath A, Banerjee A, Luo Y, Rogers D, Brooks AN, Zhu J, Haussler D. Visualizing and interpreting cancer genomics data via the Xena platform. Nat Biotechnol. 2020 Jun;38(6):675–678. doi: 10.1038/s41587-020-0546-8. PMID: 32444850; PMCID: PMC7386072.

24. Yihang Xiao, Jinyi Liu, Yan Zheng, Xiaohan Xie, Jianye Hao, Mingzhi Li, Ruitao Wang, Fei Ni, Yuxiao Li, Jintian Luo, Shaoqing Jiao, Jiajie Peng. CellAgent: An LLM-driven Multi-Agent Framework for Automated Single-cell Data Analysis. BioRxiv [Preprint] 2020 Jun 30. doi: 10.1101/2024.05.13.593861

25. Suresh S, Misra SM. Large Language Models in Pediatric Education: Current Uses and Future Potential. Pediatrics. 2024 Sep 1;154(3):e2023064683. doi: 10.1542/peds.2023-064683. PMID: 39108227.

26. Carlà MM, Gambini G, Giannuzzi F, Boselli F, De Luca L, Rizzo S. Testing the Reliability of ChatGPT Assistance for Surgical Choices in Challenging Glaucoma Cases. J Pers Med. 2025 Feb 28;15(3):97. doi: 10.3390/jpm15030097. PMID: 40137413; PMCID: PMC11943350.

27. Velez-Arce A, Li MM, Gao W, Lin X, Huang K, Fu T, Pentelute BL, Kellis M, Zitnik M. Signals in the Cells: Multimodal and Contextualized Machine Learning Foundations for Therapeutics. bioRxiv [Preprint]. 2024 Nov 12:2024.06.12.598655. doi: 10.1101/2024.06.12.598655. PMID: 38948789; PMCID: PMC11212894.

28. Zhou J, Zhang B, Li G, Chen X, Li H, Xu X, Chen S, He W, Xu C, Liu L, Gao X. An AI Agent for Fully Automated Multi-Omic Analyses. Adv Sci (Weinh). 2024 Nov;11(44):e2407094. doi: 10.1002/advs.202407094. Epub 2024 Oct 3. PMID: 39361263; PMCID: PMC11600294.

29. Bhinder B, Gilvary C, Madhukar NS, Elemento O. Artificial Intelligence in Cancer Research and Precision Medicine. Cancer Discov. 2021 Apr;11(4):900–915. doi: 10.1158/2159-8290.CD-21-0090. PMID: 33811123; PMCID: PMC8034385.

30. Chen ZH, Lin L, Wu CF, Li CF, Xu RH, Sun Y. Artificial intelligence for assisting cancer diagnosis and treatment in the era of precision medicine. Cancer Commun (Lond). 2021 Nov;41(11):1100–1115. doi: 10.1002/cac2.12215. Epub 2021 Oct 6. PMID: 34613667; PMCID: PMC8626610.

31. Zhang Z, Wei X. Artificial intelligence-assisted selection and efficacy prediction of antineoplastic strategies for precision cancer therapy. Semin Cancer Biol. 2023 May;90:57–72. doi: 10.1016/j.semcancer.2023.02.005. Epub 2023 Feb 14. PMID: 36796530.

32. Shimizu H, Nakayama KI. Artificial intelligence in oncology. Cancer Sci. 2020 May;111(5):1452–1460. doi: 10.1111/cas.14377. Epub 2020 Mar 21. PMID: 32133724; PMCID: PMC7226189.

33. Chua IS, Gaziel-Yablowitz M, Korach ZT, Kehl KL, Levitan NA, Arriaga YE, Jackson GP, Bates DW, Hassett M. Artificial intelligence in oncology: Path to implementation. Cancer Med. 2021 Jun;10(12):4138–4149. doi: 10.1002/cam4.3935. Epub 2021 May 7. PMID: 33960708; PMCID: PMC8209596.

34. Bhalla S, Laganà A. Artificial Intelligence for Precision Oncology. Adv Exp Med Biol. 2022;1361:249–268. doi: 10.1007/978-3-030-91836-1_14. PMID: 35230693.

35. Monge C, Waldrup B, Carranza FG, Velazquez-Villarreal E. Molecular Heterogeneity in Early-Onset Colorectal Cancer: Pathway-Specific Insights in High-Risk Populations. Cancers (Basel). 2025 Apr 15;17(8):1325. doi: 10.3390/cancers17081325. PMID: 40282501; PMCID: PMC12026214.

